# SARS-CoV-2 activates lung epithelia cell proinflammatory signaling and leads to immune dysregulation in COVID-19 patients by single-cell sequencing

**DOI:** 10.1101/2020.05.08.20096024

**Authors:** Huarong Chen, Weixin Liu, Dabin Liu, Liuyang Zhao, Jun Yu

## Abstract

**Objective:** The outbreak of Coronavirus disease 2019 (COVID-19) caused by SARS-CoV-2 infection has become a global health emergency. We aim to decipher SARS-CoV-2 infected cell types, the consequent host immune response and their interplay in the lung of COVID-19 patients.

**Design:** We analyzed single-cell RNA sequencing (scRNA-seq) data of lung samples from 17 subjects (6 severe COVID-19 patients, 3 mild patients who recovered and 8 healthy controls). The expression of SARS-CoV-2 receptors (ACE2 and TMPRSS2) was examined among different cell types in the lung. The immune cells infiltration patterns, their gene expression profiles, and the interplay of immune cells and SARS-CoV-2 target cells were further investigated.

**Results:** Compared to healthy controls, the overall ACE2 (receptor of SARS-CoV-2) expression was significantly higher in lung epithelial cells of COVID-19 patients, in particular in ciliated cell, club cell and basal cell. Comparative transcriptome analysis of these lung epithelial cells of COVID-19 patients and healthy controls identified that SARS-CoV-2 infection activated pro-inflammatory signaling including interferon pathway and cytokine signaling. Moreover, we identified dysregulation of immune response in patients with COVID-19. In severe COVID-19 patients, significantly higher neutrophil, but lower T and NK cells in lung were observed along with markedly increased cytokines (CCL2, CCL3, CCL4, CCL7, CCL3L1 and CCL4L2) compared with healthy controls as well as mild patients who recovered. The cytotoxic phenotypes were shown in lung T and NK cells of severe patients as evidenced by enhanced ifnγ, Granulysin, Granzyme B and Perforin expression. Moreover, SARS-CoV-2 infection altered the community interplay of lung epithelial cells and immune cells: the interaction between epithelial cells with macrophage, T and NK cell was stronger, but their interaction with neutrophils was lost in COVID-19 patients compared to healthy controls.

**Conclusions:** SARS-CoV-2 infection activates pro-inflammatory signaling in lung epithelial cells expressing ACE2 and causes dysregulation of immune response to release more pro-inflammatory cytokines. Moreover, SARS-CoV-2 infection breaks the interplay of lung epithelial cells and immune cells.

## Introduction

The Coronavirus disease 2019 (COVID-19) pandemic, caused by severe acute respiratory syndrome coronavirus 2 (SARS-CoV-2), poses a tremendous global challenge recently. As of May 7, 2020, a total of 3,820,869 COVID-19 cases and 265,098 COVID-19 deaths have been reported affecting 212 countries and territories, and the number is still growing as a result of human-to-human transmission^1,2^. SARS-CoV-2 belongs to coronaviruses family which are single-stranded and positive-sense RNA viruses characterized by club-like spike on their surface^3^. SARS-CoV-2 binds to the surface expressed proteins, angiotensin-converting enzyme 2 (ACE2), to entry into cells which is similar as SARS-CoV^4-6^ In addition to ACE2, the expression of serine protease TMPRSS2 on target cells is required for activation of viral spike (S) proteins to facilitate viral entry^6^. However, the ACE2- and TMPRSS2-expressing cell types and their expression level in the lung of COVID-19 patients are unclear.

Although SARS-CoV-2 could be recognized by the host immune system to mount an antiviral response^5,7^, imbalanced immune responses have been observed in most patients, as exemplified by high neutrophil to lymphocyte ratio^8-12^. Moreover, a large number of severe COVID-19 patients suffered cytokine storm with markedly release of proinflammatory cytokines such as interleukin 6 (IL-6), interleukin 10 (IL-10) and tumor necrosis factor (TNF)-α, leading to the progression of acute respiratory distress syndrome (ARDS) and potentially death^9,13^. However, it is still unknown how SARS-CoV-2 infection contributes to dysregulated immune response in the lung of COVID-19 patients.

In this study, we comprehensively evaluated the single cell sequencing data from the lung of 17 subjects (6 severe COVID-19 patients, 3 recovered COVID-19 patients with mild symptoms and 8 healthy donors) to uncovered cell types with ACE2 and TMPRSS2 expression in the lung infected with SARS-CoV-2. We further investigated the patterns of immune cells infiltration and their expression profiles across the different severity of infected patients as compare with the healthy controls. We finally evaluated the interactions between the gene expression profiles of lung epithelia cells and immune cells. Our investigations unravel the potential mechanisms underlying the role of SARS-CoV-2 infection in inducing the aberrant lung epithelial cell gene expression profiles and the dysregulated host immune response and their correlation changes.

## Result

### Lung epithelial cells express higher ACE2 in COVID-19 patients

To examine the expression of SARS-CoV-2 entry genes, ACE2 and TMPRSS2, in different cell types of human lung after SARS-CoV-2 infection, single-cell RNA sequencing (scRNA-seq) data of lung bronchoalveolar lavage fluid (BALF) from 3 recovered mild cases and 6 severe cases (GSE145926)^14^, as well as 8 normal lungs (GSE122960)^15^ were retrieved from NCBI database. Unsupervised analysis identified 20 distinct cell clusters (**Figure 1A and Figure S1-S3**), including epithelial (EPCAM+) and immune (PTPRC+) cell populations (**Figure S4**). ACE2 and TMPRSS2 were primarily expressed in lung epithelial cells (**Figure 1B**), in line with other studies^16,17^. Among lung epithelial populations, a relative high percentage of ACE2 or TMPRSS2 positive cells were shown in club, basal and ciliated cells which may act as primary target cells of SARS-CoV-2 infection (**Figure 1C**). Notably, the percentages of ACE2 positive cells among these three types of lung epithelial cells were all significantly higher in BALF samples from either severe or mild COVID-19 patients as compared to those in lung derived from healthy controls (**Figure 1D**). In keeping with this, the ACE2 mRNA expression level was significantly higher in COVID-19 patients compared to healthy controls in club, basal and ciliated cells, respectively (**Figure 1E**).

**Figure 1.**
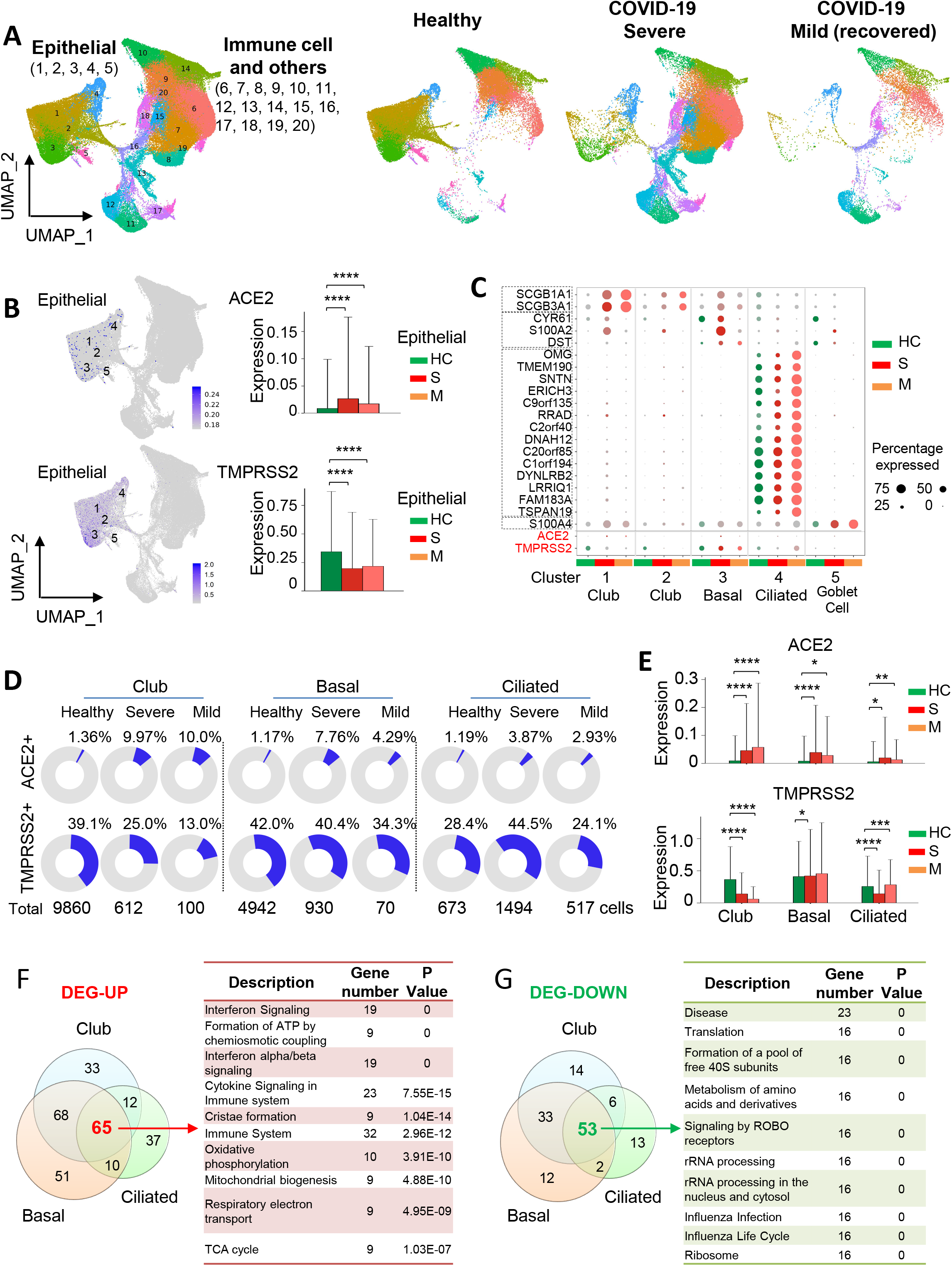
High ACE2 and TMPRSS2 expression in lung epithelial cells from COVID-19 patients. **(A)** The UMAP plot displayed the major cell types (epithelial, immune and others) in 20 clusters for bronchoalveolar lavage fluid (BALF) samples from 6 severe (S) and 3 recovered mild COVID-19 patients (M), as well as 8 healthy lung controls (HC). **(B)** UMAP plot displayed RNA expression of ACE2 or TMPRSS2. Right panel shows ACE2 or TMPRSS2 expression in lung epithelial cells from different groups. **(C)** Dot plot of ACE2 or TMPRSS2 expression for each cell-type of lung epithelial cells from different groups. Dot size represents the percentage of cells in individual clusters expressing a given gene. **(D)** The pie chart shows the percentages of ACE2- or TMPRSS2- positive cells in club (cluster 1), basal (cluster 3) and ciliated (cluster 4) cells. **(E)** Expression values of ACE2 or TMPRSS2 in different cell types of lung epithelial cells from different group. **(F and G)** Venn diagram showed overlaps among up-regulated (F) or down-regulated (G) genes in different cell-type of lung epithelial cells after SARS-CoV-2 infection (severe vs. health). Right table showed the top 10 enriched signaling pathways of common up-regulated (F) or down-regulated (G) genes. * *P* < 0.05; ** *P* < 0.01; *** *P* < 0.001; **** *P* < 0.0001 [Mann Whitney test (E)].

However, the correlation between increased ACE2 expression in the lung epithelia cells from COVID-19 patients and SARS-CoV-2 infection needs further in-depth investigation, considering the small sample size in this study and the treatment administrated to these patients.

### SARS-CoV-2 leads to cellular transcriptome alterations in lung epithelial cells

We next investigated cellular transcriptome alterations of these lung epithelial cells in response to SARS-CoV-2 infection given that they were susceptible to SARS-CoV-2 infection. Profoundly altered gene transcriptional expressions in club, basal and ciliated cells were present in COVID-19 patients (**Figure S5-S7**). A total of 65 common up-regulated genes and 53 down-regulated genes were identified in these three type of cells after virus infection (adjusted p ≤ 0.01 and |log2Fold change (FC)| ≥ 1) (**Figure 1F and 1G**). Gene Set Enrichment Analysis (GSEA) of these candidate genes revealed that SARS-CoV-2 infection induced interferon pathway and cytokine signaling in the lung epithelia cells of COVID-19 patients (**Figure 1F**). On the other hand, SARS-CoV-2 was capable to suppress host protein translation (**Figure 1G**).

### SARS-CoV-2 infection drives lung immune response

We further studied the specification of immune cells fates in response to SARS-CoV-2 infection. As shown in **Figure 1B and Figure 2A**, ACE2 or TMPRSS2 was almost not expressed in the immune cells of lung samples from COVID-19 patients, as well as healthy controls, implying that the immune cells were not susceptible to SARS-CoV-2 infection. By comparing different immune cell populations between COVID-19 patients and healthy controls, we found a dysregulated immune response in the lungs after SARS-CoV-2 infection (**Figure S8**). A massive increase of neutrophils was observed in severe COVID-19 patients compared with healthy controls, while it was restored to normal after the patients recovered (**Figure 2B**). Whilst, macrophage number was significantly lower in severe COVID-19 patients compared to healthy controls, but restored in recovered patients (**Figure 2B**). SARS-CoV-2 infection significantly increased T/NK cells in the lung, but to a lesser extent in severe COVID-19 patients (**Figure 2B**).

**Figure 2.**
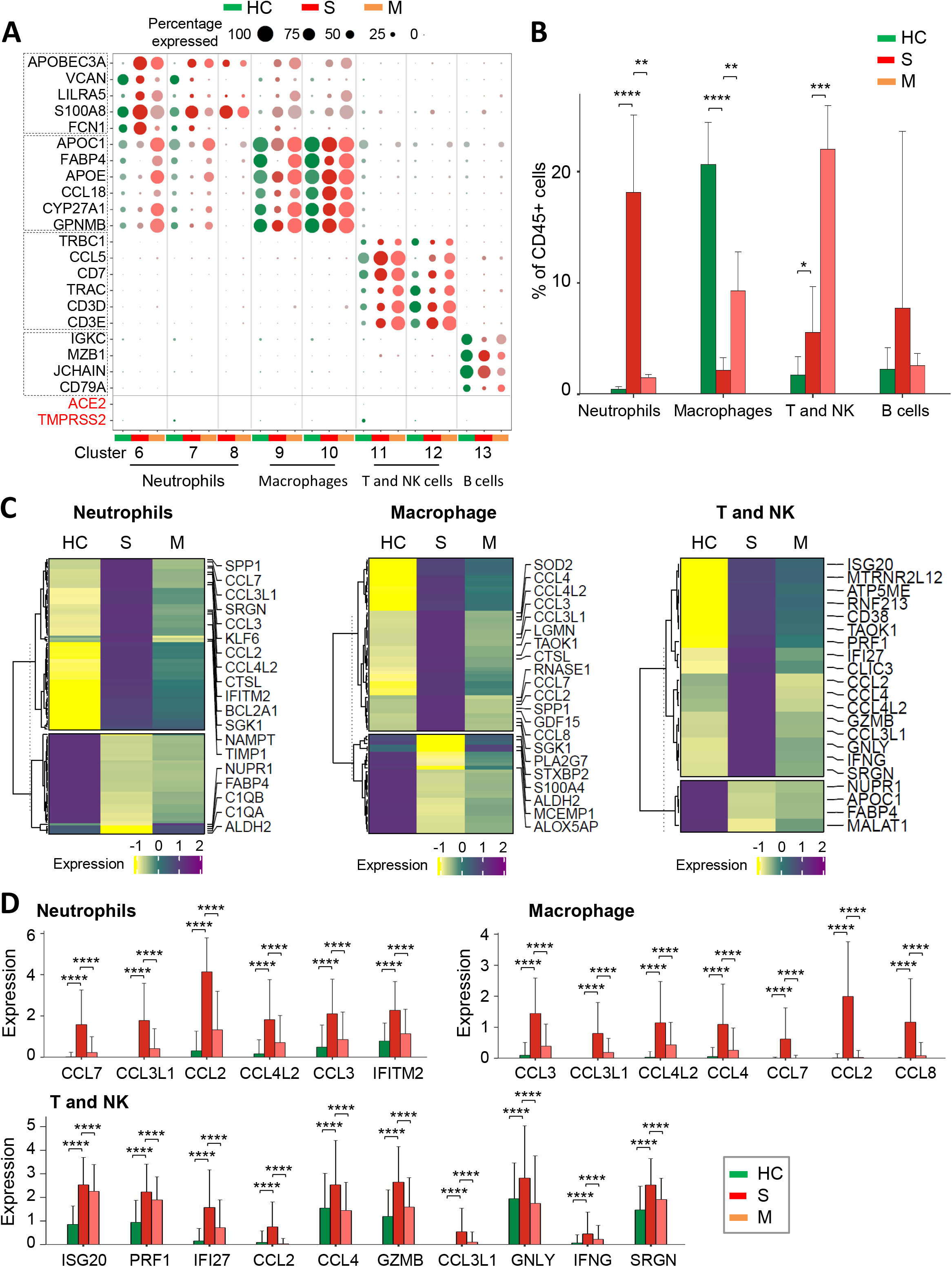
SARS-CoV-2 infection induced imbalanced host immune response in severe COVID-19 patients. **(A)** Dot plot of ACE2 or TMPRSS2 expression for each cell-type of lung immune cells from different groups. Dot size represents the percentage of cells in individual clusters expressing a given gene. **(B)** The percentages of different immune cell types of all CD45+ cells in lung samples. **(C)** Heatmaps of transcript level of candidate genes in neutrophils (cluster 7), macrophage (cluster 10) and T/NK cells (cluster 11) from different groups. **(D)** Comparison of candidate genes expression in different groups. * *P* < 0.05; ** *P* < 0.01; *** *P* < 0.001; **** *P* < 0.0001 [Mann Whitney test (B and D)].

We then explored the differential gene expression profiling of immune cells in the lung between COVID-19 patients and healthy controls. As shown in **Figure 2C**, differential gene expression patterns of neutrophil, macrogphage and T/NK cells were demonstrated in severe COVID-19 patients, mild recovered COVID-19 patients and healthy controls. In severe COVID-19 patients, we identified a variety of cytokines which were markedly increased in neutrophil, macrophage, and T/NK cells (CCL2 and CCL3L1), in neutrophil and macrophage (CCL3, CCL4L2 and CCL7), and in macrophage and T/NK cells (CCL4) respectively (**Figure 2D**). The concomitant high expression of these cytokines derived from the dysregulated immune cells attracted by SARS-CoV-2 infection suggested the occurrence of cytokine storm. Moreover, the expression of interferon Gamma (IFNG), granulysin GNLY (GNLY), granzyme B (GZMB) and perforin (PRF1) was significantly higher in T/NK cells in severe COVID-19 patients compared with healthy controls, but their expression levels were restored in the recovered patients (**Figure 2D**), suggesting that the cytotoxic T cells and NK cells were activated in response to SARS-CoV-2, and that delayed expansion of T/NK cell repertoire might present after virus infection.

### The correlation of lung epithelia cells and immune cells is changed in COVID-19 patients

We evaluated the relationship between epithelia cells and immune cells in the lung from healthy to the diseased status (**Figure 3A and 3B**). In severe COVID-19 patients, the strength of predicted strong correlations between lung epithelial cells (club and basal cells) and neutrophils were significantly reduced, but their correlations with macrophage and T/NK cell were markedly increased. As to ciliated cells, the correlation network appeared to be the same between severe COVID-19 patients and healthy control (**Figure S9**). We investigated the function of genes constituting the abovementioned network by GSEA gene enrichment analysis and found that these genes were enriched in pathways including phagosome, antigen processing, presentation and interferon alpha/beta signaling (**Figure 3C**). ASS1, CXCL8 and HLA-B were selected for further validation as they are among the list of differentially expressed genes (DEGs) in lung epithelia cells after SARS-CoV-2 infection. In both club and basal cells, the expression of ASS1, CXCL8 and HLA-B were all significantly higher in severe COVID-19 patients as compared to healthy controls or mild recovered patients (**Figure 3D**). In supporting this, SARS-CoV-2 infection significantly increased the mRNA expression of ASS1, CXCL8 and HLA-B in human normal bronchial epithelial cells, human lung cancer cell line Calu-3 and A549 overexpressing ACE2 (**Figure 3E**). Our findings suggest that the specific networks between epithelia cells and immune cells were formed in lung after SARS-CoV-2 infection.

**Figure 3.**
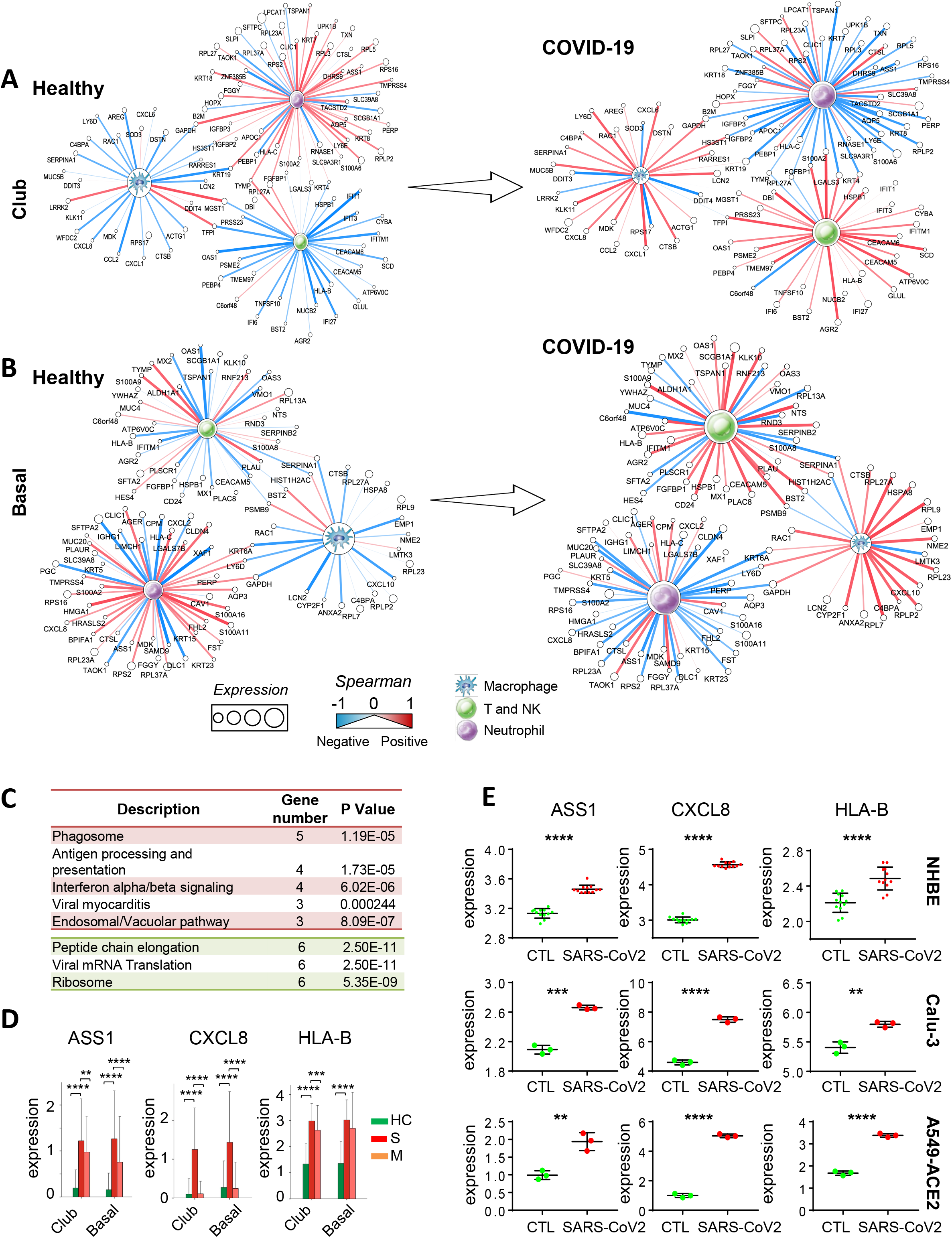
DEGs of SARS-CoV-2 target cells associated with variation of immune response. **(A)** The associations between SARS-CoV-2-induced DEGs expression in club (cluster 1) cells and frequency of immune cells (neutrophils (cluster 7), macrophage (cluster 10) and T/NK cells (cluster 11)) in health control and severe COVID-19 patients. **(B)** The associations between SARS-CoV-2-induced DEGs expression in basal (cluster 3) cells and frequency of immune cells (neutrophils (cluster 7), macrophage (cluster 10) and T/NK cells (cluster 11)) in health control and severe COVID-19 patients. **(C)** Top enriched signaling pathways of candidate genes among SARS-CoV-2-induced DEGs that were correlated with abnormal immune composition in COVID-19 patients. **(D)** Comparison of ASS1, CXCL8 or HLA-B expressions in lung epithelia cells among different groups. **(E)** Comparison of ASS1, CXCL8 or HLA-B expressions in NHBE, Calu-3 and A549-ACE2 cell lines with or without SARS-CoV2 infection. * *P* < 0.05; ** *P* < 0.01; *** *P* < 0.001; **** *P* < 0.0001 [Mann Whitney test (D) and two tailed t-test (E)].

### Discussion

In this study, we first identified high expressions of ACE2 and TMPRSS2 in ciliated, club and basal cells of lung epithelium in COVID-19 patients. ACE2 and TMPRSS2 are two critical entry genes required for SARS-CoV-2 infection^6^. The expression of ACE2 has been reported in a variety of tissues including respiratory tract and gastrointestinal mucosa^18^. ACE2 and TMPRSS2 mRNA were expressed in lung type II pneumocytes, ileal absorptive enterocytes, and nasal goblet secretory cells^17^, and their protein was detected in nasal and bronchial epithelium^19^. However, expression patterns of ACE2 and TMPRSS2 in the lung epithelial cells in COVID-19 patients have not been explored yet. This study provides the novel information on the cell types of lung epithelia cells expressing ACE2 and TMPRSS2, and their expression levels in COVID-19 patients based on single cell sequencing analyses. However, the expression of ACE2 and TMPRSS2 was not found in immune cells in the lung of COVID-19 patients. In keeping this, no SARS-CoV-2 viral gene expression was detected in peripheral blood mononuclear cell in three SARS-CoV-2 patients^20^. Thus, lung epithelium cells (ciliated, club and basal cells) but not immune cells are susceptible to SARS-CoV-2 infection.

Our further analysis demonstrated that SARS-CoV-2 led to host cellular transcriptome alterations in club, basal and ciliated cells of COVID-19 patients, resulting in activation of interferon pathway and cytokine signaling. These secreted signaling molecules serve to initiate host immune response by recruiting various immune cells^21^. In severe COVID-19 patients, the accumulated immune cells (neutrophils, macrophage and T/NK cells), especially massive infiltration of neutrophils, expressed significantly higher levels of cytokines including CCL2, CCL3, CCL4, CCL7, CCL3L1 and CCL4L2, which might contribute to lung damage. Although T cells and NK cells were reduced in the lung of severe COVID-19 patients, cytotoxic phenotypes of these cells were recognized due to significant induction of IFNG, GNLY, GZMB and PRF1. Activation of these genes were found in cytotoxic T cells (CD8+) and NK cells that inhibit virus propagation and triggers apoptosis of infected cells^22,23^. Accordingly, a higher ratio of severe COVID-19 patients was reported to present T cells specific to the viral antigen with production of infγ or GZMB in peripheral blood compared to mild COVID-19 cases^10,24^. All these findings implied that SARS-CoV-2 infection induces the aberrant epithelial cell gene expression, enriched pro-inflammation signalings and the dysregulated host immune response in lung.

By investigating the correlation between gene expression profiles of lung epithelia cells and host immune response, we discovered that the correlations between host cellular response and immune cell frequencies were altered after SARS-CoV-2 infection. The interplay between lung epithelia cells and neutrophils infiltration were diminished; in contrast, their interactions with macrophage and T/NK cell were stronger. Of these genes that were involved in the network, ASS1, CXCL8 and HLA-B expression were promoted directly by SARS-CoV-2. ASS1 ablation was reported to ameliorate liver injury by reducing neutrophil infiltration^25^. Meanwhile, CXCL8 could recruit neutrophils to the site of damage or infection.

Both CXCL8 and ASS1 presented a weaker interaction with neutrophils after SARS-CoV-2 infection, implying possible imbalanced control of innate immune response. HLA-B plays a critical role in the immune system through displaying foreign peptides. The interaction between HLA-B and T/NK cells were enhanced in the lung of COVID-19 patients, suggesting the potential activation of these immune cells by SARS-CoV-2 infection. Collectively, these data suggested that SARS-CoV-2 infection alters the interplay of lung epithelial cells and immune cells.

In conclusion, SARS-CoV-2 infection induces aberrant gene expression profiling and activation of pro-inflammatory signaling of lung epithelium cells (ciliated, club and basal cells) that expressing high levels of ACE2 and TMPRSS2. Moreover, SARS-CoV-2 infection causes dysregulated lung immune response and massive production of pro-inflammatory cytokines and disrupts the interplay of epithelial cells and immune cells. All these contribute to the consequent extensive lung damage (Figure 4).

**Figure 4.**
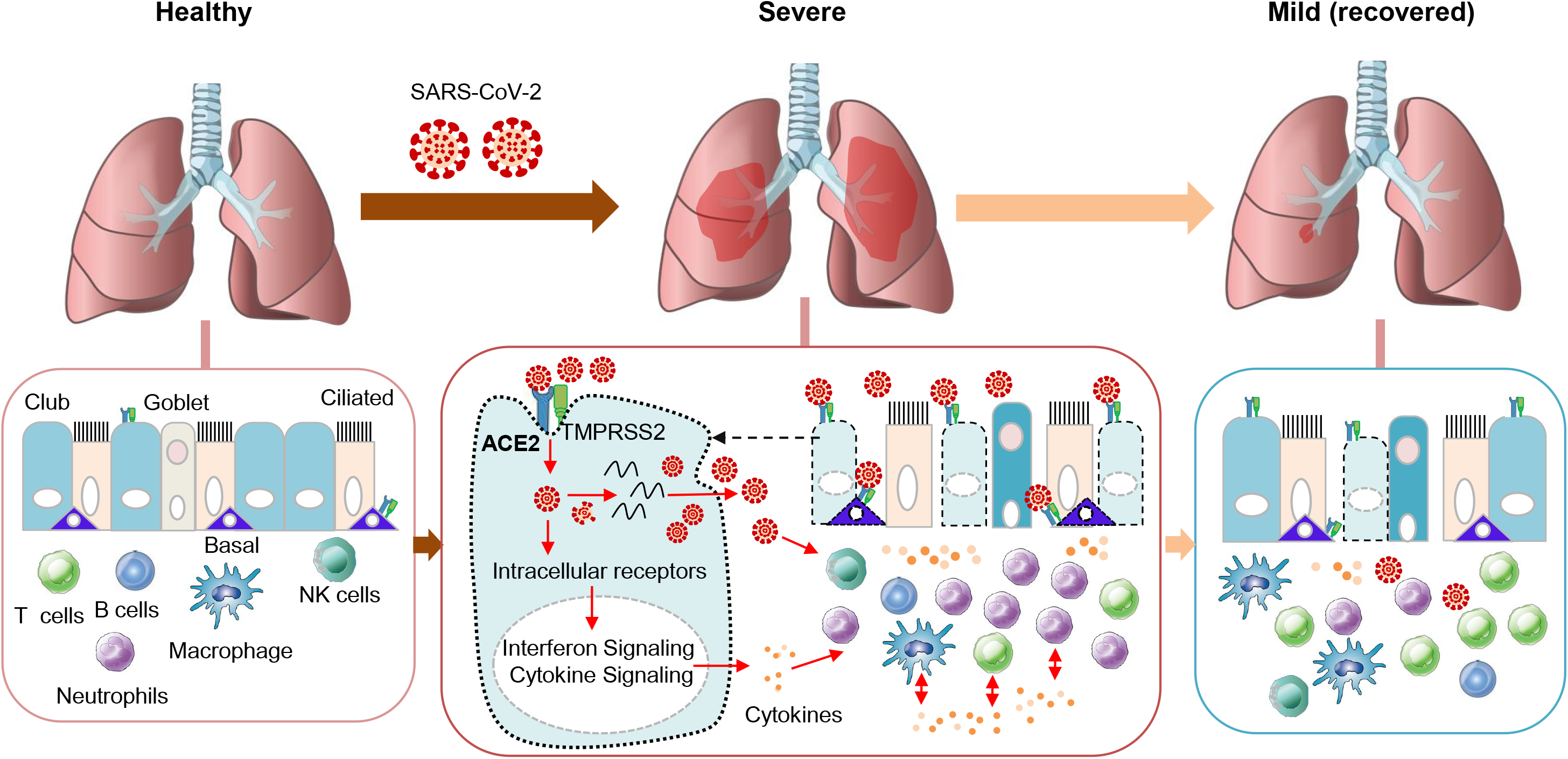
SARS-CoV-2 infection and host immune response in COVID-19 patients. In COVID-19 patients, the SARS-CoV-2 may infect ciliated cells, club cells, and basal cells expressing ACE2 and TMPRSS2 in lung epithelium and actively replicate in host cells. This could lead to activation of pro-inflammatory signaling and production of pro-inflammatory cytokines which subsequently attract both innate and adaptive immune cells including neutrophils, macrophages and T cells to the infection site to fight virus and virus-infected cells. Besides, the immune cells also release cytokines to attract more immune cells, creating a positive feedback loop of cytokine creation. Massive accumulation of pro-inflammatory cytokines producing-immune cells in the lungs could increase the severity of COVID-19 patients.

## Materials and Methods

### Datasets

Single cell RNA sequence (scRNA-Seq) data were retrieved from published resources, including bronchoalveolar lavage fluid (BALF) from 6 severe and 3 moderate COVID-19 patients^14^, and lung tissues from 8 healthy transplant donors^15^. Bulk RNA-Seq data in three SARS-CoV-2 treated cell lines were also obtained for validation purpose, including primary human bronchial epithelial cells (NHBE), Calu-3 and A549-ACE2 (with vector expressing human ACE2)^26^. All relevant data were downloaded from Gene Expression Omnibus under the accession number GSE122960, GSE145926 and GSE147507.

### scRNA-Seq data analysis

We re-analyzed the data from a count quantification matrix due to the unavailable per-cell annotation. Cells with mitochondrial gene proportion higher than 15% were filtered out. For each individual dataset, raw count matrix was first normalized and the top 2 000 most variable genes were chosen. For each cell, we divided the gene counts by the total counts and multiplied by 10 000, followed by natural-log transformation. High variable genes were determined using FindVariableFeatures in Seurat pipeline^27^. Multiple datasets were then integrated via searching the “anchors” among them^27^, enabling us to explore shared cell types presented across different datasets and conditions. The integrated data were scaled followed by principal component analysis (PCA), we retained the top 30 principal components (PCs) for further analysis. The ranking of PCs based on the variance explained by each PC were draw by ElbowPlot in Seurat and the majority of true signal is captured in the first 20 PCs (**Figure S1**). To visualize the cells, we applied the Uniform Manifold Approximation and Projection (UMAP) on the top 20 PCs. The selected PCs were also used for computing nearest-neighbour graphs and for clustering the cells. To re-annotate the cells, we identified the conserved markers for each cluster across different conditions, by comparing with all remaining clusters using FindConservedMarkers method in Seurat pipeline. Then, a manual curated cell type marker list was applied to annotate the cell type for each cluster. MAST ^28^ algorithm was used to identify the altered genes under SARS-CoV-2 infection for the epithelial-related (EPCAM+) and immune-related (CD45+) clusters, respectively.

### Bulk RNA-Seq data analysis

RNA-seq reads were mapped onto the human reference (GRCh38 with gene annotations GENCODE v30) by HISAT2 (version 2.1.0) with the default options. The number of reads mapped to each of genes was counted by using featureCount (version v1.6.4). Gene expression levels were calculated as FPKM (Fragments per Kilobase of transcript per Million mapped reads) by rpkm method in edgeR. Differentially expressed genes (DEGs) were determined using DESeq2.

### Interplay between altered genes in epithelial cells and immune cells composition

The relative amount of each immune cell type (Neutrophil, Macrophages, T/NK cells and B cells) was defined as its proportion in CD45+ cells. Correlation between gene expression and immune composition was measured by Spearman Correlation Coefficient (SCC) and was computed for healthy control and COVID-19 patients, respectively. Those with difference in SCC higher than a threshold (0.9) between healthy control and COVID-19 were chosen as differentially correlated pairs.

### Functional analysis

Functional enrichment analysis was a method aimed to identify classes of molecules (genes or proteins) that were over-represented in a set of pre-defined molecules and predicted its association with disease phenotypes. We performed this method to uncover potential biological function shift under SARS-CoV-2 infection through mapping the molecules into known molecule sets by WebGestalt ^29^. Two databases, KEGG and Reactome were used for canonical pathway detection. Significantly enriched functions were chosen if the corresponding adjusted p-value was below a threshold (0.05).

### Statistical analysis

Gene expression levels were represented as mean ± SD unless otherwise indicated. To compare the difference among groups, pairwise Wilcoxon rank sum tests was performed. All statistical analysis was conducted under R computing software.

## Data Availability

All relevant data were available in Gene Expression Omnibus under the accession number GSE122960, GSE145926 and GSE147507.

## Disclosures

The authors declared no conflict of interest.

## Author Contributions

HRC and WXL designed the study; WXL and HRC performed data analysis; HRC, WXL, DBL and LYZ contributed to the preparation of the manuscript; JY designed, supervised the study and revised the paper.

## Grant Support

This project was supported by Science and Technology Program Grant Shenzhen (JCYJ20170413161534162), HMRF Hong Kong (17160862), RGC-CRF Hong Kong (C4039-19G), RGC-GRF Hong Kong (14163817), Vice-Chancellor’s Discretionary Fund CUHK and CUHK direct grant, Shenzhen Virtual University Park Support Scheme to CUHK Shenzhen Research Institute.

## Acknowledgments

We thank Yifei Wang, Jia Yang, Shanshan Gao and Feixue Wang for their comments and suggestions for this study.

## Notes

### Competing Interest Statement

The authors have declared no competing interest.

